# MAESTRO: A Public, Generalizable Model for Stroke Lesion Segmentation from T1 MRI Across the Recovery Continuum

**DOI:** 10.64898/2026.08.22.26361044

**Authors:** Mahir H Khan, Octavio Marin-Pardo, Stuti Chakraborty, Kaitlin Lee, Sarah Lee, Nandita Raman, Juan Eugenio Iglesias, Sook-Lei Liew

## Abstract

Accurate stroke lesion segmentation is essential for large-scale neuroimaging studies, yet manual delineation remains labor-intensive, and existing automated methods often struggle to generalize across imaging protocols and stages of recovery. We developed MAESTRO, a deep learning framework for automated lesion segmentation across the stroke recovery continuum using T1-weighted (T1) MRI alone. We hypothesized that combining a transformer-based architecture with an image augmentation strategy would improve segmentation accuracy and robustness under heterogeneous imaging conditions.

T1 MRI scans and expert-traced lesion masks from 955 stroke participants across 33 international cohorts were used to train and evaluate MAESTRO within the open-source nnU-Net framework. Performance was evaluated on a held-out test set using spatial and volumetric agreement metrics. An exploratory human-in-the-loop (HITL) evaluation compared correction of MAESTRO-generated segmentations with manual tracing from scratch.

MAESTRO achieved the strongest performance across several evaluated model configurations, providing the most accurate lesion localization and lesion volume estimates (median Dice = 0.686; Pearson *r* = 0.861; ICC = 0.792). Segmentation performance was sensitive to lesion size and stroke chronicity but remained robust across diverse imaging conditions. Additionally, using a HITL workflow to correct MAESTRO segmentations reduced annotation time by 47.4% compared to manual tracing while improving accuracy relative to both automated and manual workflows.

MAESTRO is publicly available to enable robust, automated stroke lesion segmentation from T1 MRI. When combined with human review and correction, MAESTRO offers a practical approach for generating standardized, high-quality lesion annotations, helping reduce a major barrier to large-scale stroke imaging studies.

## 1. Introduction

Large neuroimaging studies have transformed stroke recovery research by providing the sample sizes needed to relate lesion location to behavioral outcomes and recovery trajectories (Bowren et al., 2022; Liew et al., 2023; Park et al., 2026; Wong et al., 2022; Zhu et al., 2010). These efforts depend on accurate delineation of focal lesions, yet manual segmentation remains a major bottleneck because it is time-consuming, resource-intensive, and difficult to scale (Tavenner et al., 2023; Ito et al., 2019). Automated lesion segmentation has the potential to substantially reduce the time required for lesion delineation, but current methods continue to face challenges in both segmentation accuracy and generalization across heterogeneous datasets (Baaklini and Valdés Hernández, 2025). As a result, human-in-the-loop (HITL) workflows that combine automated segmentation with manual review and correction may provide a practical balance between annotation efficiency and segmentation accuracy.

The potential benefits of a HITL workflow depend on the quality of the initial automated segmentation, making both accuracy and robustness across diverse imaging conditions critical. Lesion appearance evolves substantially throughout recovery, requiring training data that capture a wide range of anatomical presentations (Bachtiar et al., 2024). Accordingly, existing segmentation methods are often tailored to specific imaging modalities and stages of stroke recovery, with acute stroke segmentation typically relying on diffusion-weighted imaging (DWI) and fluid-attenuated inversion recovery (FLAIR), and chronic stroke segmentation commonly performed using T1-weighted (T1) MRI (Baaklini and Valdés Hernández, 2025; Gheibi et al., 2023; Moon et al., 2022; Shang et al., 2025; Verma et al., 2022). The availability of these imaging modalities varies across stroke stages and study cohorts (Liew et al., 2022b), further complicating efforts to develop broadly applicable segmentation models.

Because T1 MRI is often available across a wide range of stroke cohorts and recovery stages, it provides a practical foundation for combining data across studies and developing models that span the recovery continuum (Liew et al., 2022b). However, combining data across cohorts and imaging centers introduces substantial variability in scanners, acquisition protocols, spatial resolution, and image quality (Bosco et al., 2023). Deep learning approaches offer one strategy for developing segmentation models that are robust to this heterogeneity by learning features across diverse imaging and anatomical presentations. Convolutional neural networks (CNNs) have dominated medical image segmentation, learning local features that are progressively integrated into broader spatial representations (Hatamizadeh et al., 2022). Newer transformer-CNN hybrids build on this approach by using attention to more directly incorporate global context (Wald et al., 2025).

This may improve robustness to the substantial variability in stroke lesion size, location, and appearance, but remains relatively underexplored in stroke lesion segmentation. Another strategy for improving robustness to this variability is image augmentation, which introduces controlled variation in anatomy, image quality, spatial resolution, and acquisition characteristics, thus exposing models to a broader range of imaging conditions during training (Chlap et al., 2021; Hussain et al., 2018). Although augmentation has improved robustness in a variety of neuroimaging applications (Billot et al., 2023; Hoffmann et al., 2024; Iglesias et al., 2021; Laso et al., 2024), its value for T1-based stroke lesion segmentation across diverse stroke stages and imaging conditions remains unclear.

In this study, we developed MAESTRO (<u>M</u>odel for <u>A</u>utomated <u>E</u>stimation of <u>Stro</u>ke Infarcts), a T1 MRI-based deep learning framework for automated lesion segmentation across the stroke recovery continuum. Designed for public use across heterogeneous datasets, MAESTRO was trained using a large-scale, multi-site cohort encompassing acute, subacute, and chronic stroke. We hypothesized that a transformer-based implementation, combined with an image augmentation strategy, would outperform traditional CNN implementations in segmentation accuracy and robustness across heterogeneous imaging conditions. We further hypothesized that sufficiently accurate automated segmentations could serve as the foundation for a human-in-the-loop (HITL) workflow that improves annotation efficiency relative to manual tracing while maintaining high-quality lesion delineation.

## 2. Methods

### 2.1. Participants

Data used in this study was compiled by the ENIGMA Stroke Recovery working group, an international research consortium that harmonizes retrospective neuroimaging and behavioral data from diverse stroke populations (Liew et al., 2022b). While primary data collection was conducted in accordance with the Declaration of Helsinki and approved by the local ethics committee at each contributing site, the ethics committee of the receiving site (University of Southern California) approved the receipt and sharing of this pooled, de-identified dataset (HS-15-00835 approved on December 11, 2015).

### 2.2. Data Acquisition and Manual Lesion Annotation

Data was derived from the ATLAS 2.1 dataset, a classic stroke lesion segmentation benchmarking dataset and an ENIGMA Stroke Recovery Working Group resource comprising T1 MRI data from 33 international cohorts (Liew et al., 2022a). This dataset reflects the heterogeneity of stroke recovery, as participants spanned a range of post-stroke stages from acute to chronic. It also encompasses a variety of scanner manufacturers, field strengths, and spatial resolutions, reflecting real-world clinical diversity. In this study, scans were categorized according to voxel volume as higher resolution (<= 1 mm³), intermediate resolution (between 1.0 and 1.2 mm³), or lower resolution (>1.2 mm³). Because certain demographic variables are not included in the public ATLAS 2.1 release, additional demographic information, such as age, sex, and days post stroke, was obtained directly from contributing ENIGMA sites.

Stroke lesion masks were included as part of ATLAS 2.1 and were manually annotated by a team of trained researchers. Details of this standardized annotation and quality-control protocol have been reported previously (Liew et al., 2022a; Tavenner et al., 2023). Briefly, each lesion mask was independently reviewed by a second expert after the initial tracing to ensure high-quality ground truth labels. An expert neurologist was consulted to resolve any challenging cases and to ensure the accurate differentiation of stroke lesions from other anatomical features or pathologies, such as white matter hyperintensities and enlarged perivascular spaces. The resulting lesion masks served as the gold standard for downstream analyses, and lesion volume was derived from these masks by summing lesion voxels and converting voxel counts to volumetric measurements using the image resolution.

For model development, subjects were partitioned into an 80% training and 20% testing split. Subjects were first stratified by post-stroke stage and lesion size to help maintain similar distributions of clinical and anatomical characteristics across the two subsets. Post-stroke stages were categorized as acute (<7 days post-stroke), early subacute (7-90 days), late subacute (91-180 days), or chronic (>180 days) according to established recommendations by the Stroke Recovery Roundtable (Boyd et al., 2017). Lesion volumes were binned by previously established cutoffs (Ito et al., 2019): small (≤2,510 mm^3^), medium (2,510 to 21,352 mm^3^), and large (>21,352 mm^3^). This stratification strategy ensured that the training and testing data remained balanced across the full spectrum of stroke presentations.

As an exploratory analysis, 10 T1 MRI scans and their corresponding expert-derived lesion masks not used for model development were selected to comprise an *annotation evaluation dataset* used to compare a fully manual annotation workflow to a HITL annotation workflow. The expert lesion masks had undergone independent quality control and served as the reference standard. Subjects were selected to form two closely matched sets based on lesion location and lesion size.

### 2.3. Data Augmentation

We generated an augmented dataset from the original T1 scans using an augmentation workflow designed to introduce variability commonly encountered in clinical neuroimaging. The pipeline introduced variation in image geometry, anatomical morphology, spatial resolution, and image intensity through affine transformations, elastic deformations, resolution modeling, and intensity perturbations. The same spatial transforms were applied to corresponding manual lesion masks with nearest-neighbor interpolation used for label resampling. In addition, 50% of augmented samples underwent random left-right flipping, with the same transformation applied to the masks. All augmented volumes were resampled to a standardized 224 × 224 × 224 grid with isotropic 1 mm voxel spacing and are referred to as augmented T1s. Five augmented T1s were generated from each original T1, and all augmentation parameters were retained as metadata for each sample. Recorded parameters included image resolution, slice thickness, affine transformation parameters (rotation, shear, scaling, and crop-center shift), nonlinear deformation settings, intensity transformation parameters (gamma correction, bias field, blur, and noise), and left-right flip status; the sampling distributions for these parameters are summarized in Supplementary Table 1. Examples of data augmentation results from an original T1 are shown in Figure 1.

**Figure 1:**
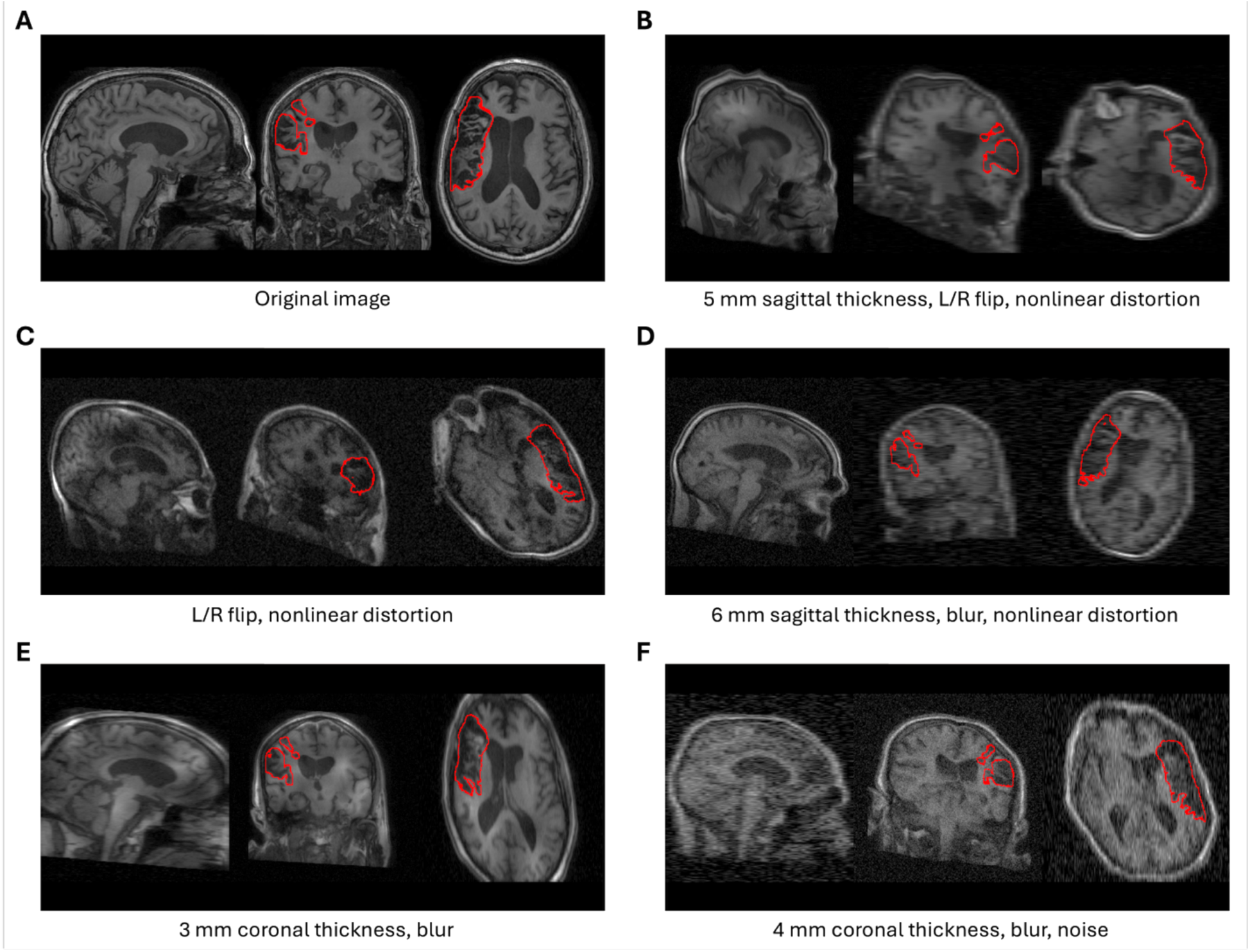
Example MRI augmentations. Panel (A) shows the original T1 MRI with lesion mask outlined in red. Augmented images were produced using randomly defined parameters from a predefined distribution to adjust image resolution, intensity blur, affine transformations, nonlinear deformation, and Gaussian noise. For this subject, augmentations included: (B) 5 mm sagittal slice thickness, left-right flip, and strong nonlinear spatial distortion; (C) left-right flip with strong nonlinear spatial distortion; (D) 6 mm sagittal slice thickness with blur and moderate nonlinear spatial distortion; (E) 3 mm coronal slice thickness with blur; (F) 4 mm coronal slice thickness with blur and Gaussian noise.

### 2.4. Model Architectures and Configurations

MAESTRO was trained using the PyTorch-based nnU-Net architecture, a publicly available self-configuring deep learning framework that automatically adapts U-Net architectures and training parameters to a given medical imaging task (Isensee et al., 2021). This design minimizes manual hyperparameter optimization and promotes standardized, reproducible performance across diverse datasets.

To evaluate the effects of both offline data augmentation and model architecture, four models were trained using combinations of the original and augmented T1 MRI datasets with two nnU-Net implementations: the standard CNN implementation and the recently introduced Primus implementation (Wald et al., 2025). Primus incorporates a hybrid transformer-convolutional architecture within the nnU-Net framework, using self-attention layers to capture broader anatomical context while retaining convolutional blocks to preserve local image detail. In contrast, the standard implementation uses a conventional residual U-Net architecture. The augmented and non-augmented Primus models were designated *MAESTRO* and *MAESTRO-NoAug*, respectively, while the corresponding standard U-Net models were designated *MAESTRO-CNN* and *MAESTRO-CNN-NoAug*.

All models used the medium-sized residual encoder planning configuration to reduce computational demands and facilitate broader reproducibility while ensuring identical preprocessing, image resampling, normalization, patch planning, and batch size selection across experiments. Images were resampled to isotropic 1 mm × 1 mm × 1 mm spacing and normalized using Z-scores. To isolate the effects of the proposed offline data augmentation strategy, nnU-Net’s default online data augmentation was disabled for all models. Model performance was evaluated using 3D full-resolution configurations.

The standard U-Net implementations were trained using the default stochastic gradient descent optimizer (initial learning rate 0.01, momentum 0.99, Nesterov acceleration enabled, weight decay 3 × 10⁻⁵), deep supervision, and a polynomial learning-rate schedule. The Primus implementations used the default Primus optimization strategy consisting of the AdamW optimizer (initial learning rate 3 × 10⁻⁴, weight decay 0.05), a warm-up learning-rate schedule followed by polynomial decay, and no deep supervision. Training followed nnU-Net’s default 5-fold cross-validation workflow, with each fold trained for 1000 epochs on NVIDIA RTX 6000 Ada Generation GPUs (48 GB VRAM).

Following model training, nnU-Net’s automated postprocessing pipeline was applied to evaluate whether postprocessing improved segmentation performance. Model selection, postprocessing, and final ensemble inference were performed using the default nnU-Net cross-validation workflow.

### 2.5. Evaluation and Statistical Analysis

#### 2.5.1. Inference on the Held-Out Test Dataset

Predicted segmentation masks were generated for all original T1 images in the held-out test set. Following nnU-Net’s standard inference procedure, predictions from the five fold-specific networks were ensembled to produce the final segmentation for each of the four model configurations (MAESTRO, MAESTRO-NoAug, MAESTRO-CNN, MAESTRO-CNN-NoAug). No image in the test set was used during model training or validation.

#### 2.5.2. Accuracy and Volumetric Agreement Assessment

We evaluated the performance of the four models by comparing their predicted lesion segmentations on the original test-set T1 images with the corresponding gold-standard manual tracings. Segmentation accuracy was quantified using the Dice similarity coefficient (DSC) to measure volumetric overlap between predicted lesion masks and manual annotations. Boundary accuracy was assessed using the average symmetric surface distance (ASSD) and the normalized surface Dice (NSD), computed using a 3 mm surface tolerance that was selected through empirical calibration of manual annotation variability (see *Supplementary Materials*). Precision and recall were also calculated to characterize the trade-off between false-positive and false-negative lesion detections. Lesion volumes were log1p-transformed prior to analysis to accommodate possible zero-volume segmentations and reduce the influence of the highly right-skewed volume distribution. Volume estimation error was defined as the absolute difference between the log-transformed gold-standard and predicted lesion volumes. Given the non-normal distribution of the performance metrics, pairwise comparisons between models were performed using paired Wilcoxon signed-rank tests. Volumetric agreement between predicted and gold-standard lesion volumes was evaluated using Pearson’s correlation coefficient and the intraclass correlation coefficient (ICC) computed on the transformed lesion volumes. ICC was calculated using a two-way random-effects model with absolute agreement for single measurements (ICC[2,1]).

#### 2.5.3. Performance Across Clinical and Lesion Characteristics

We assessed the robustness of each model across clinically relevant subgroups of the held-out test set. For each model separately, accuracy metrics (DSC, ASSD, NSD) were compared across lesion-volume categories (small, medium, and large), post-stroke stages (acute, early subacute, late subacute, and chronic), and scan-resolution categories (higher-, intermediate-, and lower-resolution) using Kruskal-Wallis tests. These analyses assessed whether overall segmentation performance varied according to lesion characteristics, stroke chronicity, or image acquisition factors. To determine whether the relative accuracy difference between models varied across subgroups, subject-level paired accuracy differences (e.g., ΔDSC = DSC[MAESTRO] - DSC[MAESTRO-NoAug]) were computed and compared across the same subgroup categories using Kruskal-Wallis tests. These analyses evaluated whether any effect of augmented-data training was consistent across clinically relevant patient and imaging.

As an exploratory analysis, we evaluated the robustness of each model to controlled image perturbations using augmented versions of the test set images. Further details can be found in the *Supplementary Materials*.

#### 2.5.4. Human-in-the-Loop Annotation Comparison

Using the annotation evaluation dataset, we conducted an exploratory analysis to compare automated, manual, and HITL annotation workflows. MAESTRO was first applied to each T1 MRI scan to generate automated lesion segmentations, which served as both the final mask for the automated workflow and the starting point for HITL annotation. Four trained lesion tracers completed manual annotation and correction of the MAESTRO-generated segmentations. Workflow assignments were balanced such that each subject underwent two independent manual annotations and two independent HITL annotations. Tracers recorded the time required to complete each annotation. Annotation time and segmentation metrics were averaged across tracers for each subject within each workflow. Because outcome measures were non-normally distributed, paired comparisons between the manual and HITL workflows were performed using Wilcoxon signed-rank tests.

## 3. Results

The final dataset included 955 stroke participants acquired across 33 sites. Of these, 761 subjects were assigned to the training set and 194 subjects were reserved as a held-out test set. Age information was available for 930 participants, among whom the mean age was 61.1 years (standard deviation [SD] = 12.9). Sex was available for 921 participants, of whom 347 (37.7%) were female. Time since stroke was available for 829 participants and had a median of 470 days (interquartile range [IQR] = 1150 days; mean [± SD] = 928.4 ± 1281.6 days). Stroke stage information was available for 934 participants, including 68 (7.3%) acute, 131 (14.0%) early subacute, 90 (9.6%) late subacute, and 645 (69.1%) chronic cases. Lesion volume had a median of 4,315 mm³ (IQR = 27,050 mm³; mean = 26,008 ± 47,944 mm³), indicating a highly right-skewed distribution of lesion sizes. The dataset comprised 793 higher-resolution (83.0%), 111 intermediate-resolution (11.6%), and 51 lower-resolution (5.3%) scans. Further details, including distributions of the training and test sets, can be found in the Supplementary Table 2.

### 3.1. MAESTRO Improves Volume Estimation While Maintaining Comparable Spatial Performance

Segmentation performance was evaluated on the original T1 images from the held-out test set (n = 194; Table 1; Supplementary Figure 4). Compared with the CNN-based models, the transformer-based models generally demonstrated superior segmentation performance (Supplementary Table 3). Relative to MAESTRO-CNN, MAESTRO achieved significantly higher NSD and recall, together with lower absolute log-transformed volume error (all FDR-adjusted *p* < 0.001), whereas MAESTRO-CNN demonstrated significantly higher precision. Although MAESTRO also demonstrated a higher median DSC than MAESTRO-CNN, this difference was not statistically significant after correction for multiple comparisons. Similarly, compared with MAESTRO-CNN-NoAug, MAESTRO-NoAug achieved significantly higher DSC, NSD, recall, and lower absolute log-transformed volume error, whereas MAESTRO-CNN-NoAug demonstrated significantly higher precision as well as lower ASSD and HD95 (all FDR-adjusted *p* < 0.05).

**Table 1.**
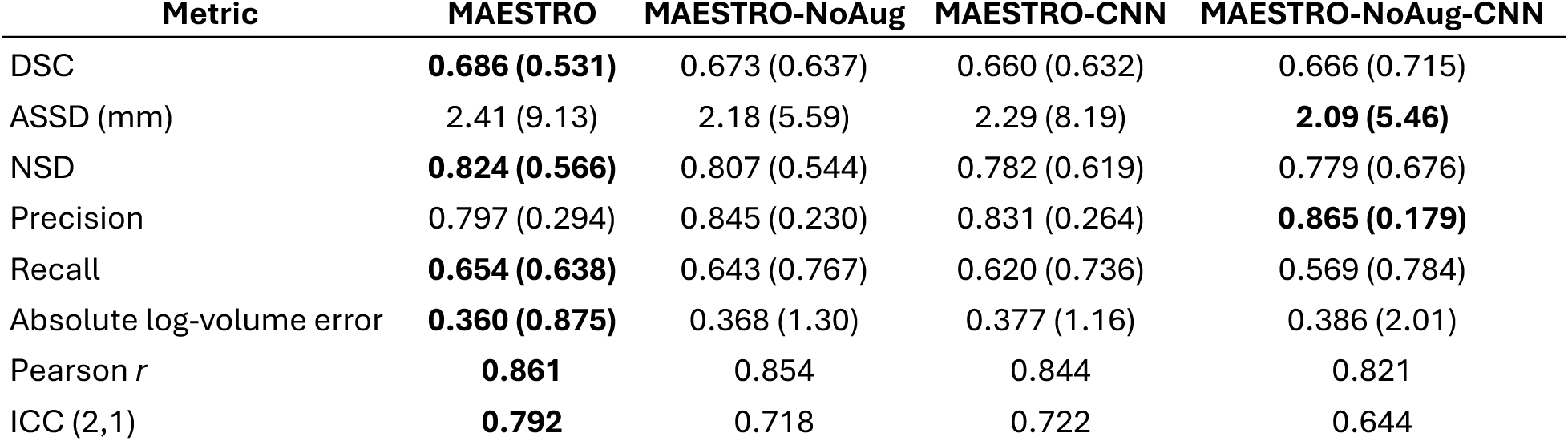
Segmentation and volumetric performance of the four models on the held-out test set. Values are reported as median (interquartile range), except for Pearson correlation coefficients and intraclass correlation coefficients (ICC), which were calculated using a two-way random-effects model with absolute agreement for single measurements (ICC[2,1]). Bold indicates the best-performing model for each metric. Abbreviations: ASSD = average symmetric surface distance; DSC = Dice similarity coefficient; ICC = intraclass correlation coefficient; NSD = normalized surface Dice.

Between models trained using transformer architectures, MAESTRO demonstrated higher median DSC, NSD, recall, and lower absolute log-transformed volume error than MAESTRO-NoAug; however, only the reduction in volume error remained statistically significant after correction for multiple comparisons (V = 6750, FDR-adjusted *p* = 0.015). Conversely, MAESTRO-NoAug demonstrated significantly higher precision than MAESTRO (V = 3841, FDR-adjusted *p* < 0.001).

Volumetric agreement between predicted and reference lesion volumes was high for all models. MAESTRO demonstrated the strongest agreement with the reference segmentations (Pearson *r* = 0.861; ICC[2,1] = 0.792), followed by MAESTRO-NoAug (*r* = 0.854; ICC[2,1] = 0.718), MAESTRO-CNN (*r* = 0.844; ICC[2,1] = 0.722), and MAESTRO-CNN-NoAug (*r* = 0.821; ICC[2,1] = 0.644).

### 3.2. Lesion Size and Post-Stroke Stage Drive Performance Variability

We next evaluated segmentation performance across lesion size, post-stroke stage, and scan type to assess model robustness across diverse stroke presentations and imaging conditions (Figure 2; Supplementary Table 4). Across all four models, segmentation performance varied significantly according to lesion size and post-stroke stage, but not scan type. DSC and NSD increased with lesion size, whereas ASSD decreased, indicating improved segmentation accuracy for larger lesions (all Kruskal-Wallis *p* < 0.05). For example, median DSC for MAESTRO increased from 0.44 for small lesions to 0.83 for large lesions, with corresponding reductions in median ASSD from 7.32 to 1.76 mm and increases in median NSD from 0.62 to 0.87. Similar trends were observed for MAESTRO-NoAug, MAESTRO-CNN, and MAESTRO-CNN-NoAug.

**Figure 2.**
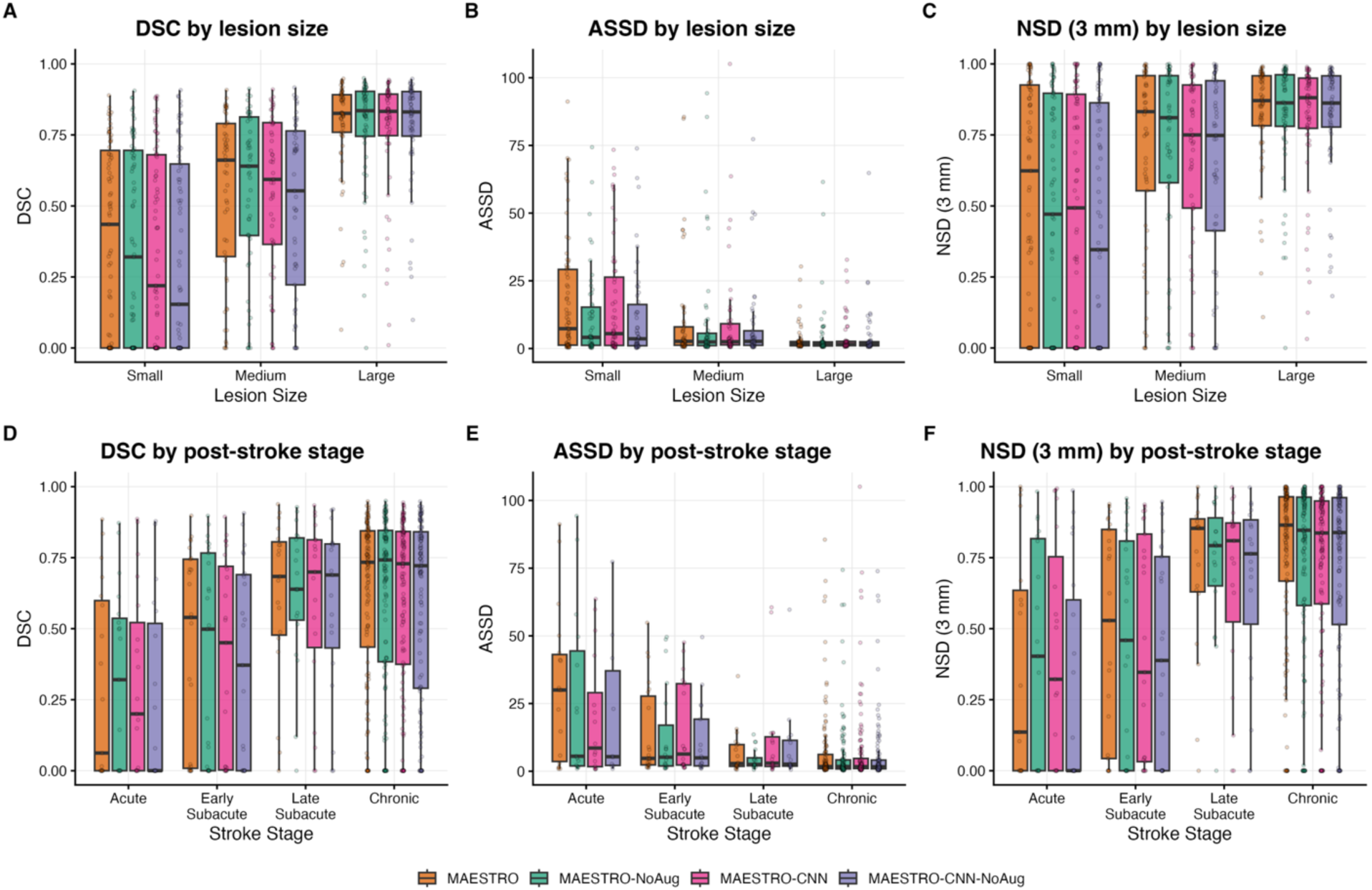
Segmentation performance stratified by lesion size and post-stroke stage. (A) DSC, (B) ASSD, and (C) NSD with 3 mm tolerance are shown across lesion size categories (small, medium, and large lesions), while (D), (E), and (F) show the same metrics across post-stroke stages (acute, early subacute, late subacute, and chronic). Boxplots show the median and interquartile range, with individual cases overlaid as points. Segmentation performance for each model improved with increasing lesion size and was highest in chronic stroke cases, with both models showing lower DSC and NSD, as well as higher ASSD for small lesions and acute-stage strokes. Although MAESTRO generally demonstrated higher performance compared to other models across subgroups, the difference in performance between models did not vary significantly by lesion size or post-stroke stage. Abbreviations: ASSD = average symmetric surface distance; DSC = Dice similarity coefficient; NSD = normalized surface Dice.

Segmentation performance also differed significantly across post-stroke stage for all models (all Kruskal-Wallis *p* ≤ 0.02). Chronic stroke cases consistently demonstrated the highest segmentation accuracy, whereas acute stroke cases exhibited the lowest performance. For MAESTRO, median DSC increased from 0.06 in acute stroke to 0.73 in chronic stroke, while median ASSD decreased from 30.0 to 1.74 mm and median NSD increased from 0.14 to 0.86. Comparable improvements across post-stroke stages were observed for the remaining models.

In contrast, segmentation performance was largely unaffected by scan type. No significant differences in DSC, ASSD, or NSD were observed across clinical, research, and borderline scan types for any model (all Kruskal-Wallis *p* > 0.18), indicating that segmentation performance was robust to variation in image acquisition.

Pairwise Kruskal-Wallis tests of the performance differences between model pairs identified no significant subgroup effects for DSC, ASSD, or NSD after FDR correction (all adjusted *p* > 0.05), indicating that the relative performance differences between models were not significantly modified by lesion size, scan type, nor stroke stage.

### 3.3. Human-in-the-Loop Annotation Reduces Annotation Time and Improves Segmentation Accuracy

In an exploratory evaluation of tracings on 10 T1 MRI scans, the HITL workflow generally reduced annotation time compared with manual tracing (Figure 3). Across all subjects, the median difference in annotation time was 11.1 minutes, representing a 47.4% reduction relative to manual tracing (V = 52, *p* = 0.01). Absolute time savings varied across cases, with the largest reduction approaching two hours. Compared with manual tracing, the HITL workflow significantly improved segmentation accuracy, with higher DSC and NSD (FDR-corrected *P* < 0.05). Median DSC values were 0.768, 0.732, and 0.709 for the HITL, manual, and automated workflows, respectively; median NSD values were 0.868, 0.829, and 0.850; and median ASSD values were 1.74, 2.34, and 1.69 mm. No other pairwise comparisons were statistically significant after FDR correction.

**Figure 3:**
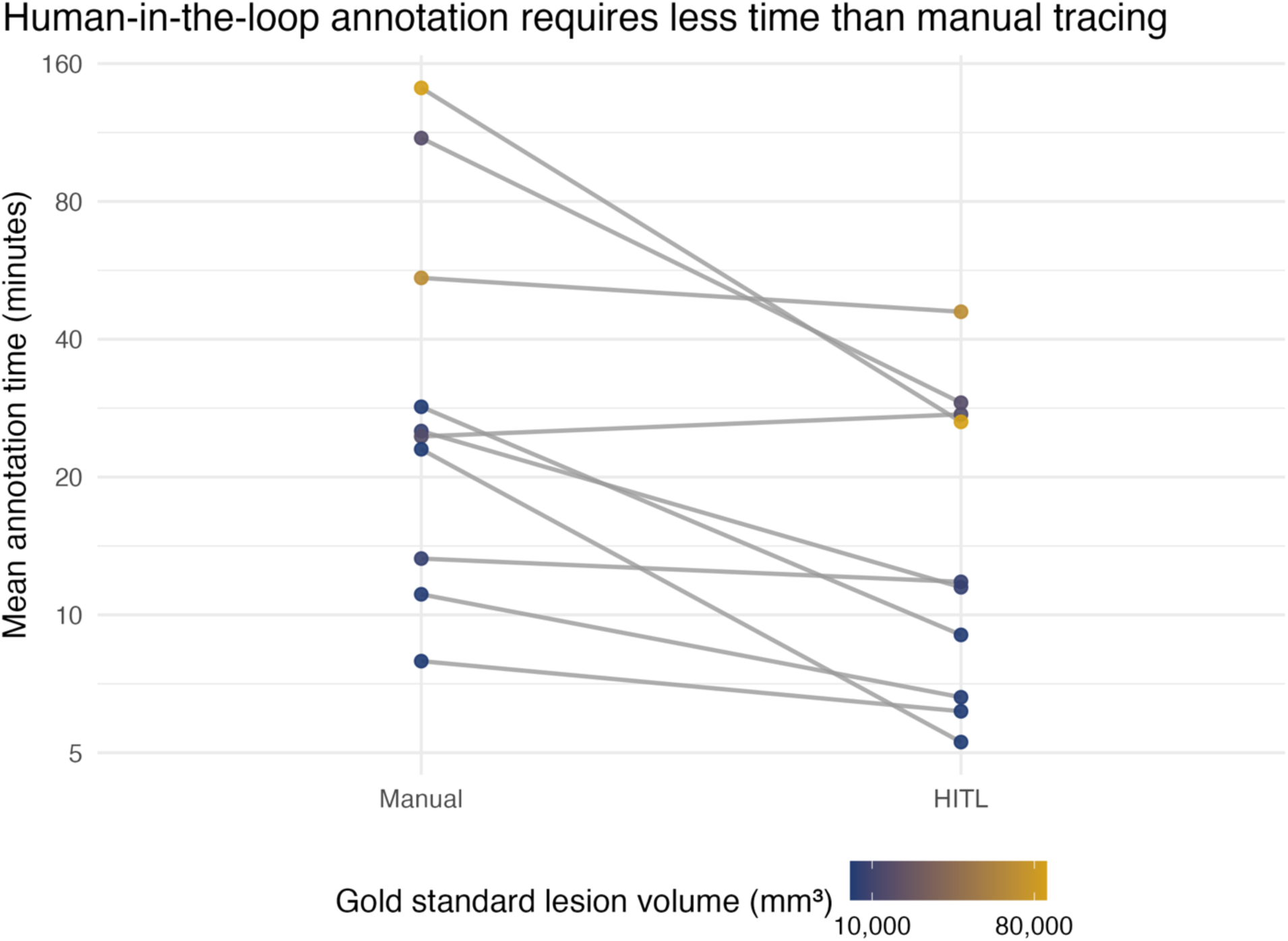
Human-in-the-loop annotation reduces annotation time compared with manual tracing. Each point represents the mean annotation time for a single subject, averaged across two independent tracers for each workflow. The y-axis is displayed on a logarithmic scale to emphasize proportional reductions in annotation time. Point color indicates lesion volume derived from the expert reference mask. Annotation time was reduced for most evaluation subjects when initialized using MAESTRO and corrected through the human-in-the-loop workflow. Larger lesions generally required more annotation time and exhibited larger absolute time reductions with human-in-the-loop correction. Abbreviations: HITL = human-in-the-loop.

## 4. Discussion

In this study, we developed MAESTRO, a T1 MRI-based automated lesion segmentation framework designed for heterogeneous, multisite stroke datasets spanning the full recovery continuum. By combining a transformer-based nnU-Net architecture with an offline image augmentation strategy that modeled real-world variation in image acquisition and quality, MAESTRO outperformed variants that used conventional CNN architectures or omitted offline augmentation, providing both the most accurate lesion localization and the most reliable lesion volume estimates. Together, these findings establish MAESTRO as a practical framework for automated lesion segmentation and lay the foundation for scalable human-in-the-loop lesion annotation, making standardized, high-quality lesion annotation feasible at a scale that was previously impractical using manual tracing alone.

Among all evaluated model configurations, MAESTRO consistently provided the strongest overall balance between spatial accuracy and lesion volume estimation. Although improvements in overlap-based metrics relative to the other transformer-based model were modest and generally consistent with previously reported T1-based stroke lesion segmentation methods (Baaklini and Valdés Hernández, 2025; Ito et al., 2019), MAESTRO achieved the lowest lesion volume estimation error and the highest agreement with expert-derived lesion volumes. This improved performance likely reflects the complementary contributions of the transformer-based architecture and the proposed offline augmentation strategy. The transformer architecture leverages broader anatomical context through self-attention while preserving local anatomical detail through convolutional components (Wald et al., 2025), which may be an advantage for stroke lesions that often exhibit heterogeneous morphology and indistinct boundaries. Meanwhile, the offline augmentation strategy exposed the model to greater variation in image quality, spatial resolution, and acquisition characteristics during training, better reflecting the diversity encountered in large multisite datasets. Consistent with this interpretation, augmentation primarily improved lesion volume estimation while also increasing robustness to simulated image perturbations, suggesting that exposure to realistic imaging variability produces more stable predictions under heterogeneous acquisition conditions.

Performance varied substantially according to both lesion size and stroke stage, two factors that have consistently challenged automated lesion segmentation methods (Baaklini and Valdés Hernández, 2025; Ito et al., 2019). Segmentation performance improved with increasing stroke chronicity, likely because lesions become more conspicuous on T1 MRI as biological changes stabilize over time (Bachtiar et al., 2024). Conversely, smaller lesions produced lower segmentation accuracy across all model configurations; this reflects in part the increasing sensitivity of overlap-based metrics such as DSC to differences in boundary placement as lesion size decreases. We therefore also evaluated normalized surface Dice, which assesses whether lesion boundaries fall within an acceptable spatial tolerance (Gut et al., 2022; Ostmeier et al., 2023). Here, we used numerous qualitatively approved lesion tracings from human raters to identify a 3-mm tolerance as the degree of boundary variation considered acceptable during manual annotation. MAESTRO demonstrated the strongest performance for both Dice similarity coefficient and normalized surface Dice at this 3-mm tolerance, suggesting improved boundary localization rather than simply greater volumetric overlap. Nevertheless, because segmentation performance remained lowest for small lesions and acute stroke, careful manual review remains particularly important in these more challenging cases.

Because manual review remains important for many lesion segmentations, particularly in more challenging cases, we assessed whether MAESTRO could serve as the foundation for a human-in-the-loop workflow that shifts the annotation process from manual lesion tracing to review and correction. Although this evaluation was exploratory and involved a relatively small number of cases, correcting MAESTRO segmentations reduced annotation time by nearly half while improving segmentation accuracy relative to both fully automated and fully manual workflows. These findings support automated segmentation and expert review as complementary components of an efficient annotation pipeline rather than competing approaches. As additional validation becomes available, we anticipate adopting this workflow as the standard lesion annotation protocol within the ENIGMA Stroke Recovery Working Group, which motivated the accompanying annotation protocol (see *Supplementary Materials*) to facilitate broader implementation and standardization across research groups.

Several limitations should be considered when interpreting these findings. Although the training dataset was designed to span the stroke recovery continuum and diverse imaging conditions, acute stroke and lower-resolution scans remained underrepresented. We attempted to mitigate these imbalances by stratifying the training and testing splits and designing the augmentation strategy to mimic lower-quality clinical acquisitions, but additional validation in independent datasets, particularly with sufficient acute stroke and lower-resolution imaging, will be important for further establishing generalizability. We also chose to use an offline augmentation strategy rather than nnU-Net’s standard online augmentation pipeline. Although manually defining augmentation parameters may introduce biases that differ from standardized online augmentation, this approach ensured identical augmented images across cross-validation folds and enabled controlled robustness analyses using the same augmentation pipeline on the held-out test data. More broadly, these findings suggest that image augmentation may have value beyond model training by providing a framework for evaluating prediction robustness under controlled imaging perturbations.

MAESTRO provides a publicly available framework for automated stroke lesion segmentation from T1 MRI across the full spectrum of stroke recovery, which can be found at https://github.com/npnl/MAESTRO. Built within the open nnU-Net ecosystem and accompanied by publicly released model weights, MAESTRO can readily incorporate future advances in segmentation architectures and training strategies while continuing to be adapted and refined as additional annotated stroke datasets become available. When integrated with human review and correction, MAESTRO provides a practical approach for generating standardized, high-quality lesion annotations across heterogeneous stroke cohorts, helping to reduce one of the major practical barriers to large-scale stroke imaging studies.

## Acknowledgements

The authors acknowledge the following individuals whose lesion tracings completed during their training contributed to a subset of the analyses presented in this work: Ashley Catchpole, Ning-Yun Chang, Rebecca Isip, Jace Kim, Amisha Kumar, Justin Lee, and Sasha Medvidovic.

During the preparation of this manuscript, the authors used ChatGPT 5.5 Instant (OpenAI) to revise the text for clarity and linguistic refinement; the authors reviewed and edited the output as needed and take full responsibility for the final content of the article.

## Data Availability

MAESTRO is publicly available at https://github.com/npnl/MAESTRO. MAESTRO was developed using nnU-Net, which is available at https://github.com/MIC-DKFZ/nnUNet. ATLAS data is available at https://fcon_1000.projects.nitrc.org/indi/retro/atlas.html.

