## Supplementary Materials for "MAESTRO: A Public, Generalizable Model for Stroke Lesion Segmentation from T1 MRI Across the Recovery Continuum"

#### Supplementary Methods

##### SM.1. Normalized Surface Dice Tolerance Calibration

The normalized surface Dice (NSD) requires specification of a surface tolerance ($\tau$), which defines the maximum distance at which two surface points are considered to agree (Nikolov et al., 2021). To determine an appropriate tolerance for this study, 10 independent raters generated manual lesion segmentations for five representative scans without access to existing annotations. All segmentations underwent qualitative review by trained reviewers and were deemed acceptable relative to an expert reference annotation. NSD was computed between each rater's segmentation and the reference annotation across surface tolerances from 1 to 5 mm (1-mm increments) to characterize the level of boundary agreement achievable by trained human raters.

##### SM.2. Robustness and Prediction Stability Analysis

We evaluated the extent to which segmentation performance remained stable under simulated variations in image quality and acquisition characteristics using the augmented test-set T1 images. Performance relative to the corresponding augmented gold-standard lesion masks was assessed using DSC, ASSD, NSD, precision, recall, and volume estimation error. For each performance metric, robustness was quantified as the within-subject coefficient of variation (CoV) across the five augmented images, with lower values indicating greater prediction stability across image perturbations. Because CoV distributions were non-normal, differences between models were evaluated using paired Wilcoxon signed-rank tests.

To determine which sources of image variability most strongly influenced model performance, separate linear mixed-effects models were fit for each performance metric, with individual augmentation parameters specified as fixed effects and subject included as a random intercept to account for repeated measurements across augmented images. Predictor and outcome variables were standardized prior to model fitting, allowing effect sizes to be compared across perturbation types and performance metrics. Statistical significance was assessed for the fixed-effect term corresponding to each augmentation parameter, with false discovery rate (FDR) correction applied to account for multiple testing.

#### Supplementary Results

##### SR.1. Normalized Surface Dice

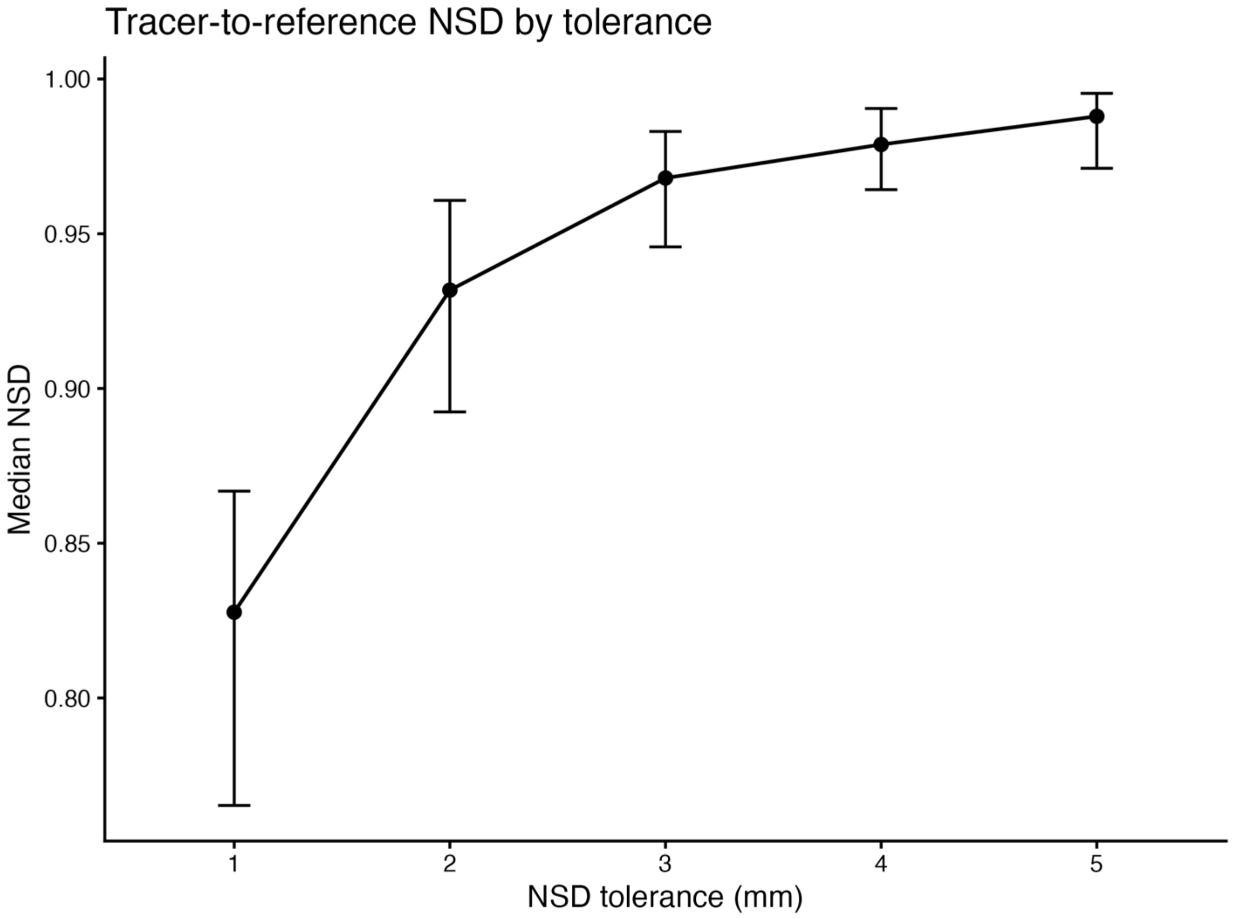


**Supplementary Figure 1. Empirical calibration of the normalized surface Dice (NSD) tolerance.** Median tracer-to-reference NSD is shown across surface tolerances from 1 to 5 mm. Error bars represent the interquartile range. Agreement increased monotonically with increasing tolerance, with the largest gains occurring between 1 and 2 mm and progressively smaller improvements thereafter.

Tracer-to-reference NSD increased monotonically with increasing surface tolerance (Supplementary Figure 1). Median NSD increased from 0.828 (IQR: 0.765-0.867) at 1 mm to 0.932 (IQR: 0.892-0.961) at 2 mm and 0.968 (IQR: 0.946-0.983) at 3 mm, with smaller increases at 4 mm (0.979, IQR: 0.964-0.990) and 5 mm (0.988, IQR: 0.971-0.995). Thus, increasing the tolerance beyond 3 mm produced relatively small gains in agreement despite requiring progressively larger surface discrepancies to be considered a match. Given the heterogeneity in spatial resolution across T1 MRI acquisitions, a 3 mm tolerance was selected for the primary analyses to accommodate expected variability in boundary placement.

#### SR.2. MAESTRO Demonstrates Greater Robustness to Image Perturbations

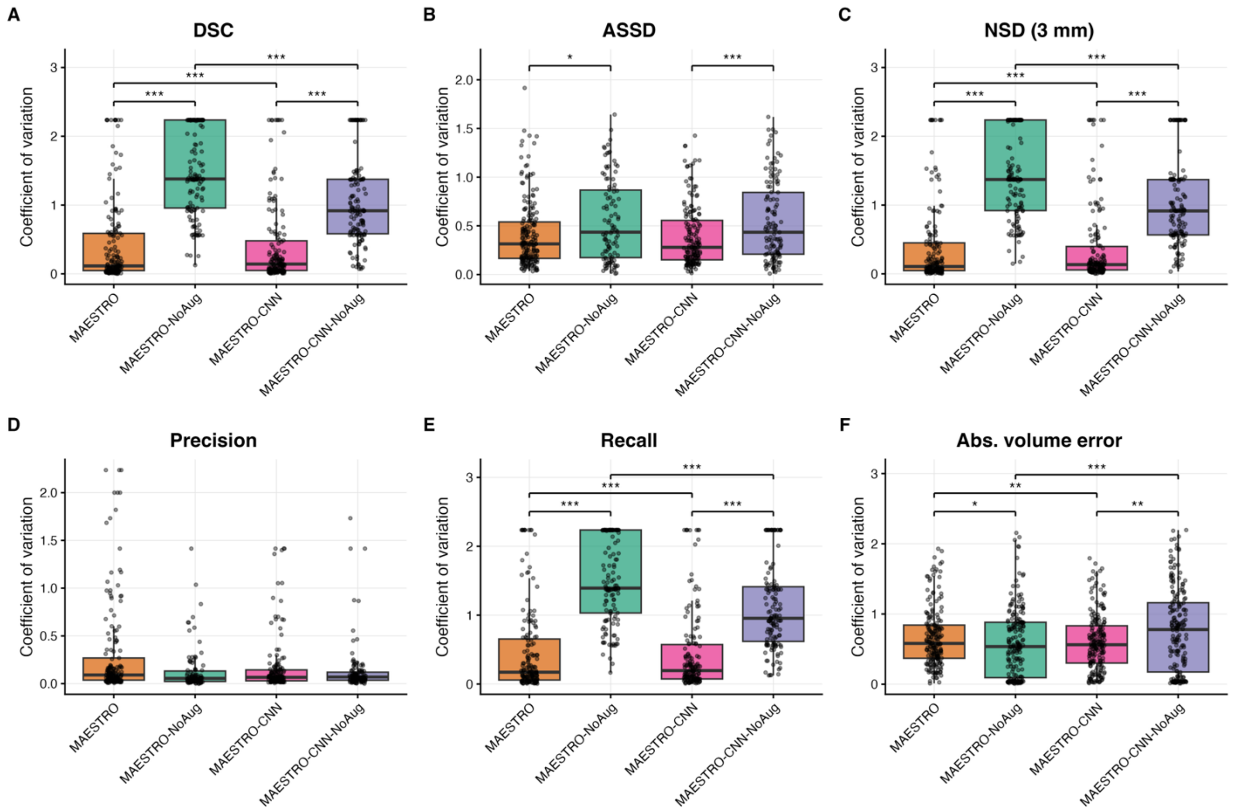


**Supplementary Figure 2. Robustness of MAESTRO and MAESTRO-NoAug to augmented image perturbations.** Robustness was quantified using the within-subject coefficient of variation (CoV) across five augmented T1 images generated for each subject, with lower CoV values indicating greater consistency. Panels show CoV for (A) DSC, (B) ASSD, (C) NSD with 3 mm tolerance, (D) precision, (E) recall, and (F) absolute log-transformed volume estimation error. Boxplots show the median and interquartile range, with individual subjects overlaid as points. Differences between models were assessed using paired Wilcoxon signed-rank tests and denoted as $P < 0.05$ (*), $P < 0.01$ (**), and $P < 0.001$ (***). Besides precision, the models trained on augmented data (MAESTRO, MAESTRO-CNN) exhibited lower variability for each metric compared to their counterpart models trained on original data (MAESTRO-NoAug, MAESTRO-CNN-NoAug). Abbreviations: ASSD = average symmetric surface distance; DSC = Dice similarity coefficient; NSD = normalized surface Dice.

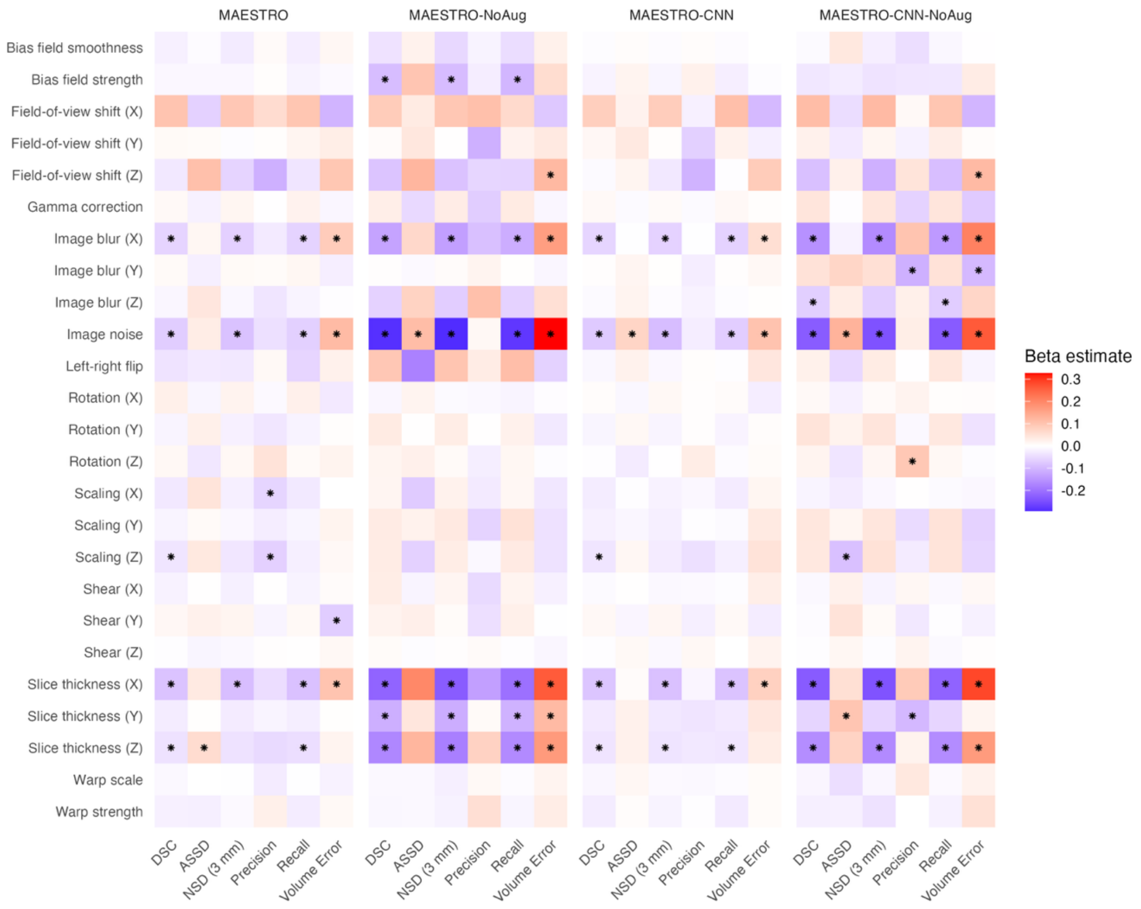


**Supplementary Figure 3. Full heatmap of associations between image perturbations and segmentation performance for each model.** Heatmaps show standardized regression coefficients (β) from linear mixed-effects models relating image perturbations to segmentation performance for each model. Color indicates the magnitude and direction of the estimated association (red = positive, blue = negative), and asterisks denote associations that remained statistically significant after multiple-comparison correction (*P*<0.05, **P*<0.01, ***P*<0.001). Rows correspond to image perturbations and columns to performance metrics (DSC, HD95, precision, recall, and volume estimation error). Across models, image blur, increased slice thickness, and image noise exhibited the strongest associations with segmentation performance, with the MAESTRO-NoAug and MAESTRO-CNN-NoAug variants generally showing a greater number of significant associations than their augmented counterparts. Abbreviations: ASSD = average symmetric surface distance; DSC = Dice similarity coefficient; NSD = normalized surface Dice.

To assess robustness to image perturbations, we calculated the within-subject coefficient of variation (CoV) across the five augmented T1 images generated for each test subject. Overall, augmentation substantially improved robustness for both architectures (Supplementary Figure 2). The augmented MAESTRO and MAESTRO-CNN models exhibited substantially lower variability than their corresponding non-augmented variants. Median

DSC CoVs were 0.115 for MAESTRO and 0.142 for MAESTRO-CNN, compared with 1.38 for MAESTRO-NoAug and 0.917 for MAESTRO-CNN-NoAug. Corresponding NSD CoVs were 0.106, 0.134, 1.37, and 0.914, respectively. Median ASSD CoVs were 0.314 and 0.279 for MAESTRO and MAESTRO-CNN, compared with 0.435 and 0.434 for their non-augmented counterparts. Precision exhibited relatively low variability across all models (median CoV, 0.055–0.091). Paired Wilcoxon signed-rank tests demonstrated significantly lower within-subject variability for the augmented compared with the corresponding non-augmented model for most segmentation metrics.

We next examined associations between image perturbations and segmentation performance to identify factors contributing to model robustness (Supplementary Figure 3). After false discovery rate (FDR) correction ($P < 0.05$), image noise and increased slice thickness along the x-axis were the most consistent predictors of performance degradation across all four models. These perturbations were associated with lower DSC, NSD, and recall, together with increased absolute volume error, with the largest effect sizes observed for the non-augmented models. For example, image noise was more strongly associated with reduced DSC in MAESTRO-NoAug ($\beta = -0.288$) and MAESTRO-CNN-NoAug ($\beta = -0.231$) than in MAESTRO ($\beta = -0.070$) and MAESTRO-CNN ($\beta = -0.075$). Similarly, associations with absolute volume error were larger for the non-augmented models ($\beta = 0.327$ and 0.263, respectively) than for their augmented counterparts ($\beta = 0.115$ and 0.102). Similarly, increased slice thickness along the x-axis was associated with reduced segmentation performance across all models but exhibited markedly larger effect sizes in the non-augmented variants. Additional significant associations were observed primarily in the non-augmented models, including sensitivity to slice thickness in the y- and z-directions and bias-field variation (MAESTRO-NoAug), whereas the augmented models showed relatively few significant associations beyond image noise and slice thickness.

### Supplementary Tables

| Parameter | Bounds |
|---|---|
| Image Resolution | Isotropic (50%): Each axis sample independently from U(1.0, 1.2) mm<br>Clinical (50%): Two axes fixed at 1.0 mm, final from U(2.5, 6) mm |
| Rotation | U(-15°, 15°) on each axis |
| Shear | U(-0.2, 0.2) on each axis |
| Scaling | U(0.8, 1.2) on each axis |
| Nonlinear deformation scale | U(0.03, 0.06) |
| Nonlinear deformation strength | U(0, 4) |
| Gamma correction | LogNormal(0.0, 0.1) |
| Bias-field scale | U(0.02, 0.04) |
| Bias-field strength | U(0.0, 0.6) |
| Noise standard deviation | U(0.0, 0.15) |
| Left-right flip | Bernoulli(p = 0.5) |

**Supplementary Table 1. Distributions for data augmentation parameters.** Parameters for each data augmentation were sampled according to the distributions shown. Crop-center shift and blur are omitted because they are derived from the input image dimensions and sampled slice thickness, respectively, rather than sampled from fixed bounded distributions.

| | Full Dataset (n=955) | Training Set (n=761) | Testing Set (n=194) |
|---|---|---|---|
| Sites Represented | 33 | 32 | 22 |
| Age (years) (mean ± SD) | 61.1 ± 12.9 | 61.1 ± 12.9 | 61.2 ± 13.0 |
| Sex (%) | | | |
| Female | 347 (37.6%) | 275 (37.4%) | 72 (38.5%) |
| Male | 573 (62.2%) | 458 (62.4%) | 115 (61.5%) |
| Time Since Stroke (days) (median [IQR]) | 470 (1150) | 457 (1145) | 541 (1167.8) |
| Stroke Stage (%) | | | |
| Acute | 68 (7.3%) | 49 (6.6%) | 19 (10.1%) |
| Early Subacute | 131 (14.0%) | 106 (14.2%) | 25 (13.3%) |
| Late Subacute | 90 (9.6%) | 71 (9.5%) | 19 (10.1%) |
| Chronic | 645 (69.1%) | 520 (69.7%) | 125 (66.5%) |
| Estimated Lesion Volume ($mm^3$) (median [IQR]) | 4,314.9 (27,050.2) | 4,367 (26,687.0) | 4,188.7 (30,242.6) |
| Lesion Size (%) | | | |
| Small (<2,510 $mm^3$) | 398 (41.7%) | 319 (41.9%) | 79 (40.7%) |
| Medium | 267 (27.9%) | 214 (28.1%) | 53 (27.3%) |
| Large (>21,352 $mm^3$) | 290 (30.4%) | 228 (30.0%) | 62 (32.0%) |
| Scan Resolution (%) | | | |
| Higher (≤ 1 $mm^3$) | 793 (83.0%) | 626 (82.3%) | 167 (86.1%) |
| Intermediate | 111 (11.6%) | 100 (13.1%) | 11 (5.7%) |
| Lower (≥ 1.2 $mm^3$) | 51 (5.3%) | 35 (4.6%) | 16 (8.2%) |

**Supplementary Table 2. Demographic, clinical, lesion, and imaging characteristics of the complete dataset and the corresponding training and held-out test sets.** Continuous variables are reported as mean ± standard deviation (SD) or median (interquartile range [IQR]) as appropriate. Percentages were calculated using subjects with available data for each variable.

| Comparison | Metric | Wilcoxon V | FDR-adjusted *p* |
|---|---|---|---|
| **MAESTRO vs. MAESTRO-CNN** | DSC | 8616 | 0.072 |
| | ASSD | 6367 | 0.123 |
| | HD95 | 5018 | 0.076 |
| | NSD | 9267 | **<0.001** |
| | Precision | 4055 | **<0.001** |
| | Recall | 10277 | **<0.001** |
| | Volume error | 5900 | **<0.001** |
| **MAESTRO-NoAug vs. MAESTRO-CNN-NoAug** | DSC | 8788 | **<0.001** |
| | ASSD | 4422 | **0.002** |
| | HD95 | 3622 | **0.012** |
| | NSD | 8762 | **<0.001** |
| | Precision | 2879 | **<0.001** |
| | Recall | 10525 | **<0.001** |
| | Volume error | 4154 | **<0.001** |
| **MAESTRO vs. MAESTRO-NoAug** | DSC | 6630 | 0.386 |
| | ASSD | 7115 | 0.916 |
| | HD95 | 5475 | 0.392 |
| | NSD | 7672 | 0.293 |
| | Precision | 3841 | **<0.001** |
| | Recall | 7989 | 0.119 |
| | Volume error | 6750 | **0.015** |
| **MAESTRO-CNN vs. MAESTRO-CNN-NoAug** | DSC | 7622 | 0.386 |
| | ASSD | 5936 | 0.856 |
| | HD95 | 4790 | 0.392 |
| | NSD | 7721 | 0.156 |
| | Precision | 3412 | **<0.001** |
| | Recall | 9575 | **<0.001** |
| | Volume error | 5583 | **<0.001** |

**Supplementary Table 3. Pairwise comparisons of segmentation and volumetric performance between models.** Pairwise differences were assessed using paired Wilcoxon signed-rank tests. P-values were adjusted for multiple comparisons using the Benjamini-Hochberg false discovery rate (FDR) procedure.

| Metric | Model | Lesion size $\chi^2$(df) | Lesion size *P* | Scan type $\chi^2$(df) | Scan type *P* | Stroke stage $\chi^2$(df) | Stroke stage *P* |
|---|---|---|---|---|---|---|---|
| DSC | MAESTRO | 68.28 (2) | **<0.001** | 2.38 (2) | 0.304 | 19.92 (3) | **<0.001** |
| | MAESTRO-NoAug | 63.21 (2) | **<0.001** | 3.33 (2) | 0.189 | 19.27 (3) | **<0.001** |
| | MAESTRO-CNN | 64.29 (2) | **<0.001** | 1.64 (2) | 0.441 | 16.79 (3) | **<0.001** |
| | MAESTRO-CNN-NoAug | 68.40 (2) | **<0.001** | 2.82 (2) | 0.244 | 22.33 (3) | **<0.001** |
| ASSD | MAESTRO | 14.06 (2) | **<0.001** | 0.36 (2) | 0.834 | 24.03 (3) | **<0.001** |
| | MAESTRO-NoAug | 7.82 (2) | **0.020** | 0.48 (2) | 0.788 | 7.82 (2) | **0.020** |
| | MAESTRO-CNN | 9.43 (2) | **0.009** | 0.94 (2) | 0.624 | 15.76 (3) | **0.001** |
| | MAESTRO-CNN-NoAug | 6.86 (2) | **0.032** | 2.46 (2) | 0.292 | 16.55 (3) | **<0.001** |
| NSD (3 mm) | MAESTRO | 13.88 (2) | **<0.001** | 0.82 (2) | 0.664 | 22.94 (3) | **<0.001** |
| | MAESTRO-NoAug | 24.71 (2) | **<0.001** | 2.49 (2) | 0.288 | 22.49 (3) | **<0.001** |
| | MAESTRO-CNN | 20.57 (2) | **<0.001** | 1.24 (2) | 0.538 | 16.47 (3) | **<0.001** |
| | MAESTRO-CNN-NoAug | 28.69 (2) | **<0.001** | 1.58 (2) | 0.454 | 27.35 (3) | **<0.001** |

**Supplementary Table 4. Kruskal-Wallis tests evaluating differences in segmentation performance across lesion size, scan type, and post-stroke stage for each MAESTRO model.** Segmentation performance varied significantly by lesion size and post-stroke stage, but not by scan type, for both models. Test statistics ($\chi^2$), degrees of freedom (df), and *P* values are reported for the Dice similarity coefficient (DSC), average symmetric surface distance (ASSD), and normalized surface Dice (NSD; 3 mm tolerance).

### Supplementary Figures

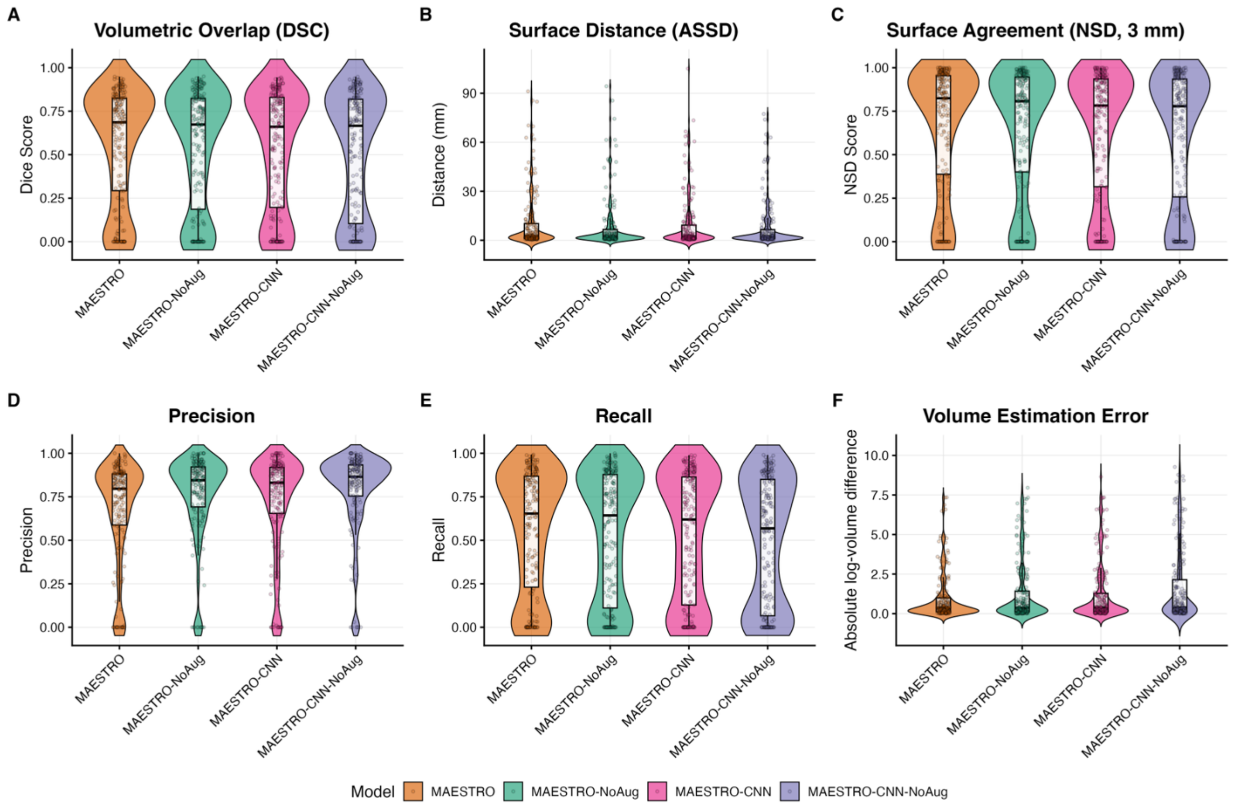


**Supplementary Figure 4. Comparative performance of MAESTRO, MAESTRO-NoAug, MAESTRO-CNN, and MAESTRO-CNN-NoAug across segmentation and volumetric evaluation metrics.** Violin plots show the distribution of performance across individual test cases, with overlaid boxplots indicating the median and interquartile range. (A) Volumetric overlap measured by the Dice similarity coefficient (DSC), with higher values indicating greater agreement between predicted and reference segmentations. (B) Surface distance measured by the average symmetric surface distance (ASSD), where lower values indicate more accurate boundary delineation. (C) Surface agreement measured by the normalized surface Dice (NSD) at a 3-mm tolerance, with higher values indicating better surface correspondence. (D) Precision, representing the proportion of predicted lesion voxels that were true positives. (E) Recall, representing the proportion of reference lesion voxels correctly identified by each model. (F) Volume estimation error, quantified as the absolute log-transformed difference between predicted and reference lesion volumes, with lower values indicating more accurate volume estimation. Overall, MAESTRO shows the strongest overall performance, with significant gains in overlap, boundary accuracy, surface agreement, recall, and volumetric accuracy, at the cost of lower precision. Abbreviations: DSC, Dice similarity coefficient; ASSD, average symmetric surface distance; NSD, normalized surface Dice.

### Supplementary Appendix

#### SA.1. Human-in-the-Loop Lesion Annotation Protocol

The results of the present study demonstrate that MAESTRO substantially reduces annotation time by providing accurate initial lesion segmentations for human review. Because segmentation performance remains less reliable for smaller lesions and other challenging cases, automated segmentation should complement rather than replace manual annotation. Based on these findings, we developed the following human-in-the-loop (HITL) lesion annotation protocol, which has been adopted by the ENIGMA Stroke Recovery Working Group as the standard workflow for lesion annotation.

##### *SA.1.1. Tracer Training*

The previous ENIGMA Stroke Recovery Working Group tracer training protocol, described by Tavenner et al. (2023), emphasized manual lesion identification and delineation. Briefly, during Phase 1, trainees manually traced lesions from five T1 MRI scans, revising their annotations until approved by a trained reviewer. After a one-week interval, trainees began Phase 2, in which they repeated the same five scans along with two additional unseen cases before becoming approved tracers.

The revised protocol retains the original Phase 1 because lesion identification and proficiency with the tracing software remain essential skills. After completing the first five approved manual tracings, trainees proceed to a revised Phase 2 consisting of 10 previously unseen scans accompanied by initial lesion segmentations. Rather than tracing lesions from scratch, trainees review and correct these segmentations until they match the expert reference masks. The training cases were selected to include common model failure modes, including small lesions, under- and over-segmentation, and confounding pathologies such as white matter hyperintensities and enlarged perivascular spaces. This phase is intended to develop the skills required for routine HITL lesion annotation.

Throughout training, trainees have access to a curated collection of reference materials describing stroke lesion identification and differentiation from common imaging confounds. These resources include written guidance and representative imaging examples that have been reviewed by neuroradiologists and experienced lesion experts and are available throughout training as reference materials.

To provide objective feedback throughout training, we developed an automated evaluation script. The script compares trainee-generated lesion masks with expert reference annotations, computes quantitative performance metrics, and generates visual reports highlighting regions of disagreement. Quantitative metrics include Dice similarity coefficient (DSC), normalized surface Dice (NSD; 1-, 2-, 3-, 4- and 5-mm tolerances), average symmetric surface distance (ASSD), 95th percentile Hausdorff distance (HD95), lesion volume, lesion count, false-positive volume, and false-negative volume. It also generates visual reports highlighting regions of disagreement, allowing trainees and reviewers to rapidly identify over- and under-segmentation. By providing standardized quantitative and visual feedback, the automated evaluation framework reduces reviewer burden while promoting consistent assessment of trainee performance.

###### *SA.1.2. Annotation Workflow*

The revised annotation workflow begins by applying MAESTRO to each T1 MRI scan to generate an initial lesion segmentation. First, a trained tracer reviews the automated segmentation and corrects any errors, including missed lesions, over-segmentation, under-segmentation, and misclassification of non-stroke pathology. The corrected segmentation is then independently reviewed by a second trained tracer, who performs any additional revisions before approving the final lesion mask. To maintain consistency across studies, a designated lesion expert provides guidance on challenging cases, resolves disagreements between reviewers, and oversees annotation standards. When lesion identity remains uncertain, consultation with neuroradiology experts is performed as needed before the lesion mask is finalized.

This workflow preserves independent multi-stage quality control while shifting tracer effort from manual lesion tracing to review and correction of automated segmentations.
